# Ethanol-Drying Regeneration of N95 Respirators

**DOI:** 10.1101/2020.04.12.20059709

**Authors:** Albert I. Nazeeri, Isaac A. Hilburn, Daw-An Wu, Kabir A. Mohammed, D. Yovan Badal, Moses H.W. Chan, Joseph L. Kirschvink

**Affiliations:** Division of Geological and Planetary Sciences, California Institute of Technology, Pasadena CA 91125, USA; Division of Biology and Biological Engineering, California Institute of Technology, Pasadena CA 91125, USA; Department of Physics, The Pennsylvania State University, University Park, Pennsylvania 16802, USA; Earth-Life Science Institute, Tokyo Institute of Technology, Meguro, 152-8550, Tokyo, Japan

**Keywords:** Laser Particle Counting, Melt-Blown Polypropylene Microfibers, Vacuum Drying, COVID-19, Personal Protective Equipment, Adsorbed Water, Electret Filter

## Abstract

A critical shortage of respirators, masks and other personal protective equipment (PPE) exists due to the COVID-19 pandemic. Of particular need are N95 respirators, which use meltblown microfibers of charged polypropylene. An intensive search is underway to find reliable methods to lengthen the useful life of these normally disposable units.

Recent experiments on respirators cleaned with ethanol solutions found drastic post-treatment drops infiltration efficiency (>40%). This has been attributed to a mechanism whereby ethanol disrupts the charges in the microfibers, reducing their ability to trap particles. The CDC/NIOSH has issued guidance directing clinicians and researchers to pursue other methods of decontamination.

In our experiments, we replicated the drop in efficiency after 70% ethanol treatment, but we found that the efficiency rose again after more effective drying, which we achieved with a vacuum chamber. After drying at pressures of < ∼6 mbar (0.6 kPa), the measured filtering efficiency rose to within 2% of the pre-washing value, and we found that this was sustained for 5 cleaning-drying cycles in three models of N95 masks. We stress that our tests are not meant to certify that the respirators are safe for use, which would require further, standardized, testing under NIOSH protocols. The tests presented here are used to understand basic mechanisms by which treatments can decrease or increase filtration efficiency.

The main mechanism underlying the loss and recovery of filter efficiency seems to be the deposition and removal of water molecules adsorbed on the fiber surfaces, a hypothesis which is supported by several observations: (A) the filtering efficiency increases non-linearly with the weight loss during drying. (B) filtration efficiency shows an abrupt recovery as the vacuum pressure drops from 13 to 6 mbar, the range physically attributable to the removal of adsorbed water. (C) Optical microscopy of the microfiber layer reveals surface wetting of the fibers, which is most resistant to drying in dense regions of the fiber network. These observations indicate that losses in filter efficiency may be caused by the wicking of water into the dense fiber networks, reducing the available surface area for filtration.

Such a degradation mechanism has two implications: (A) Ethanol and other aqueous decontamination methods may be more viable than previously assumed. Investigations of such methods should specify drying methods in their protocols. We employ vacuum chambers in this study, but other methods of removing adsorbed water could be equivalent. (B) This mechanism presents the possibility that mask filtration performance may be subject to degradation by other sources of moisture, and that the mask would continue to be compromised even if it appears dry. Further research is needed to determine the conditions under which such risks apply, and whether drying should be a routine practice for respirators undergoing extended use.

This study introduces a number of methods which could be developed and validated for use in resource-limited settings. As the pandemic continues to spread in rural areas and developing nations, these would allow for local efforts to decontaminate, restore, and test medical masks.

## Introduction

Medical respirators and masks are in critically short supply across the globe, in particular the N95 and equivalent designations. Most of these masks rely on charged, melt-blown polypropylene microfibers and are designed to be disposable (1). During use, the filters may become contaminated with viral or bacterial particles, necessitating that any efforts to refurbish them employ decontamination techniques.

A simple broad-spectrum method for decontamination is to rinse or soak materials in 60-80% ethanol, which is a potent virucidal agent against lipophilic viruses (3). Unfortunately, recent studies (6, 11) reported that the performance of N95 respirators degrades by as much as ∼40% after immersion in a solution of 70-75% alcohol. Previous studies (9, 11) postulated a degradation mechanism whereby penetration of alcohol into mask polypropylene microfibers permanently disrupts surface electrical charges, reducing the ability of the fibers to trap aerosols. The CDC/NIOSH have issued guidance directing clinicians and researchers to pursue other forms of decontamination (4, 12).

We report here experiments that raise a different mechanism: filtration efficiency appears to be hampered by a film of water molecules that remains adsorbed on the microfiber surfaces. Due to the fibers’ large surface area, approximately 2-5 g of water remains adsorbed on the fibers inside a typical respirator even after extensive air drying. This water-adsorption-based degradation is reversible by drying, which we demonstrate using a vacuum chamber to incrementally draw out different fractions of the solution.

## Methods

### Measurement of Medical Respirator and Mask Efficiency

Prior to the pandemic we routinely monitored the quantity of particulates in a magnetically-shielded, biomagnetic clean lab (10) in the 0.3, 0.5, 1.0, 5.0 and 10 μm size bins using a MetOne Aerocet 531S laser particle counter. These units sample air at ambient pressure with a small pump, running it through the laser detectors and counting particles in five size bins. In order to use this to sample the air passing through a filter, we constructed a testing rig as shown in Fig. 1. We wrapped a Styrofoam bust of a human head in three layers of ordinary kitchen plastic wrap to give surface friction and pliancy, drilled a vent hole “mouth” with a hot copper pipe, and placed the N95 respirators over the opening. This setup was housed inside a Plexiglass box, which was then pressurized with ‘dirty’ Pasadena air (the pressurized box reached ∼2,000,000 particles ≥ 0.3 μm/cf) using an industrial heat gun dryer (a Gilson MA-290F) in non-heat mode, mounted to the side. Particulates passing through the apparatus include those in the laboratory air, as well as any generated by the hardware, such as the heat gun used as a blower. Air passed through the filters and then exited through a PVC pipe, which was sampled by the particle counter at ambient pressure. The air then flowed through a 2 m length of plastic hose to minimize backflow of outside air into the counter from turbulence. The flow rate through the mask was adjusted using the intake manifold on the heat gun to ∼23 l/min.

**Figure 1:**
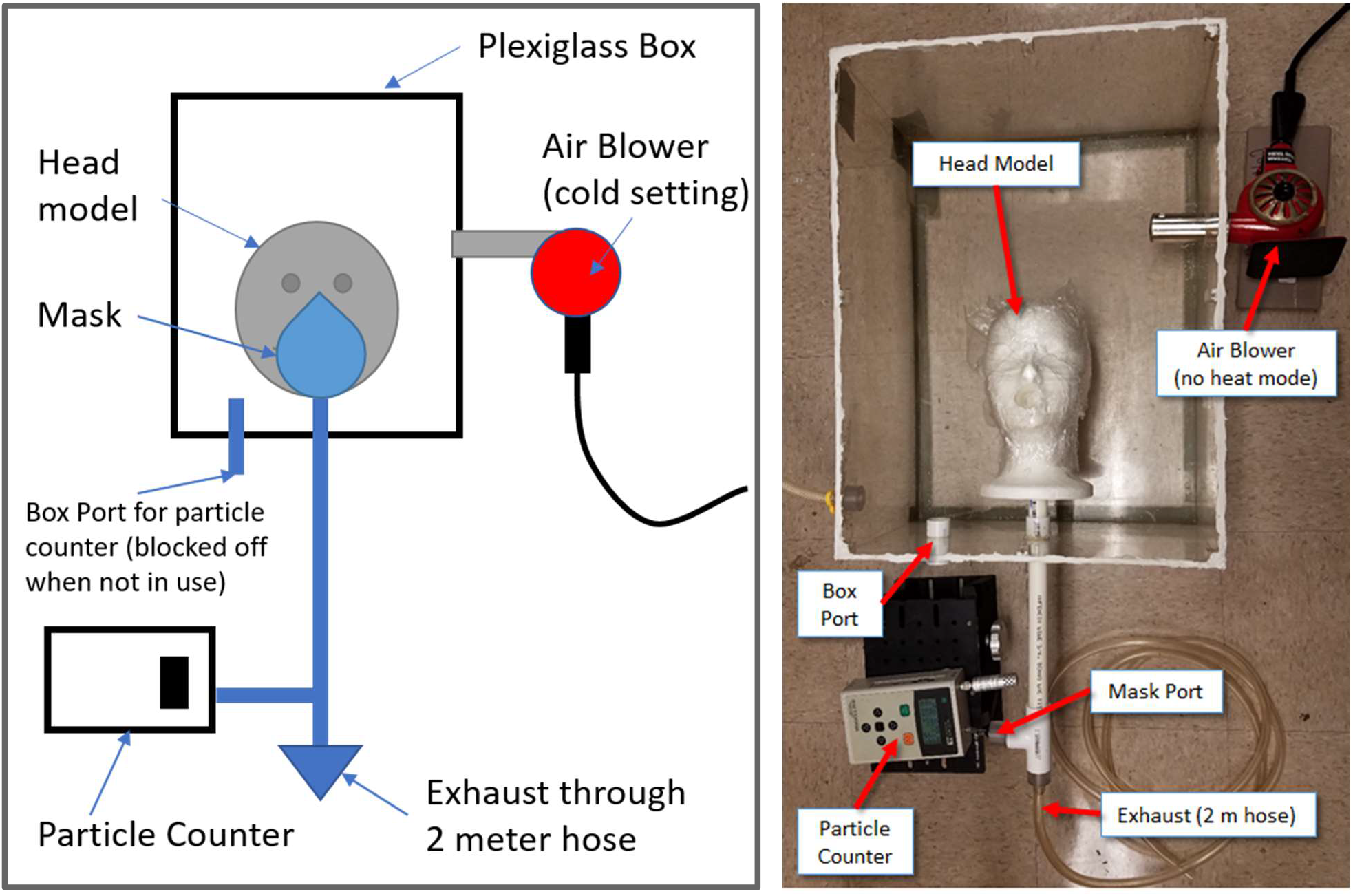
Left, schematic diagram of the experimental medical filter testing chamber. Right: Image of the current system with the top removed. The hand-held air blower forces ambient dirty air (∼2 million particles at and above 0.3 μm/cuf) into the chamber, which exits either through the mask/head plumbing system or through the background port. The ratio of the particle counts at and above 0.3 μm between the two ports gives a relative measure of the mask efficiency.

We found that minor deviations in flow rate had no noticeable effect on the efficiency measure. Every two minutes the particle counter inlet was switched between the mask port (post-filtration) and a box port (unfiltered background air in the box), and the ratios of the total particle counts were used to calculate the filtration efficiency. The data were transmitted via USB serial link and stored to file on a local computer. This computer system both collected data from the particle counter and controlled the device via custom-built software in C#, adapted from software developed for monitoring air quality in the aforementioned clean lab (10).

### Mask Cleaning Protocol

The 70% v/v ethanol solution was prepared with 200 proof laboratory grade ethanol and deionized (DI) water, which is within the CDC guidelines for disinfection (3). Approximately 50 mL of ethanol solution was poured over each mask, such that every part of the mask was saturated. Excess liquid was blotted off with a paper towel, and the masks were allowed to air dry for 2-3 hours before the vacuum experiments were conducted.

### Data Analysis

Filtration efficiencies were calculated from the ratios of the raw incoming and outgoing counts from the laser particle counter in the ≥0.3 μm size fraction. The error in the filtration efficiency was split into two different types: 1) error introduced by variations in the fit of the mask, and 2) the counting error of the particle fluxes. Error from the fit was determined by refitting the mask three to four times and calculating the standard deviation from the mean of the measurements. Error in the measured fluxes was found by taking two to three measurements in steady state per mask fitting and calculating the standard deviation from the mean of the measurements. The total error in the filtration efficiency was found through quadrature. Error in the masks’ masses (Fig. 2) was found through the difference in mask mass before and after the efficiency measurement (this difference is due to some of the adsorbed water being removed by airflow during testing).

**Figure 2:**
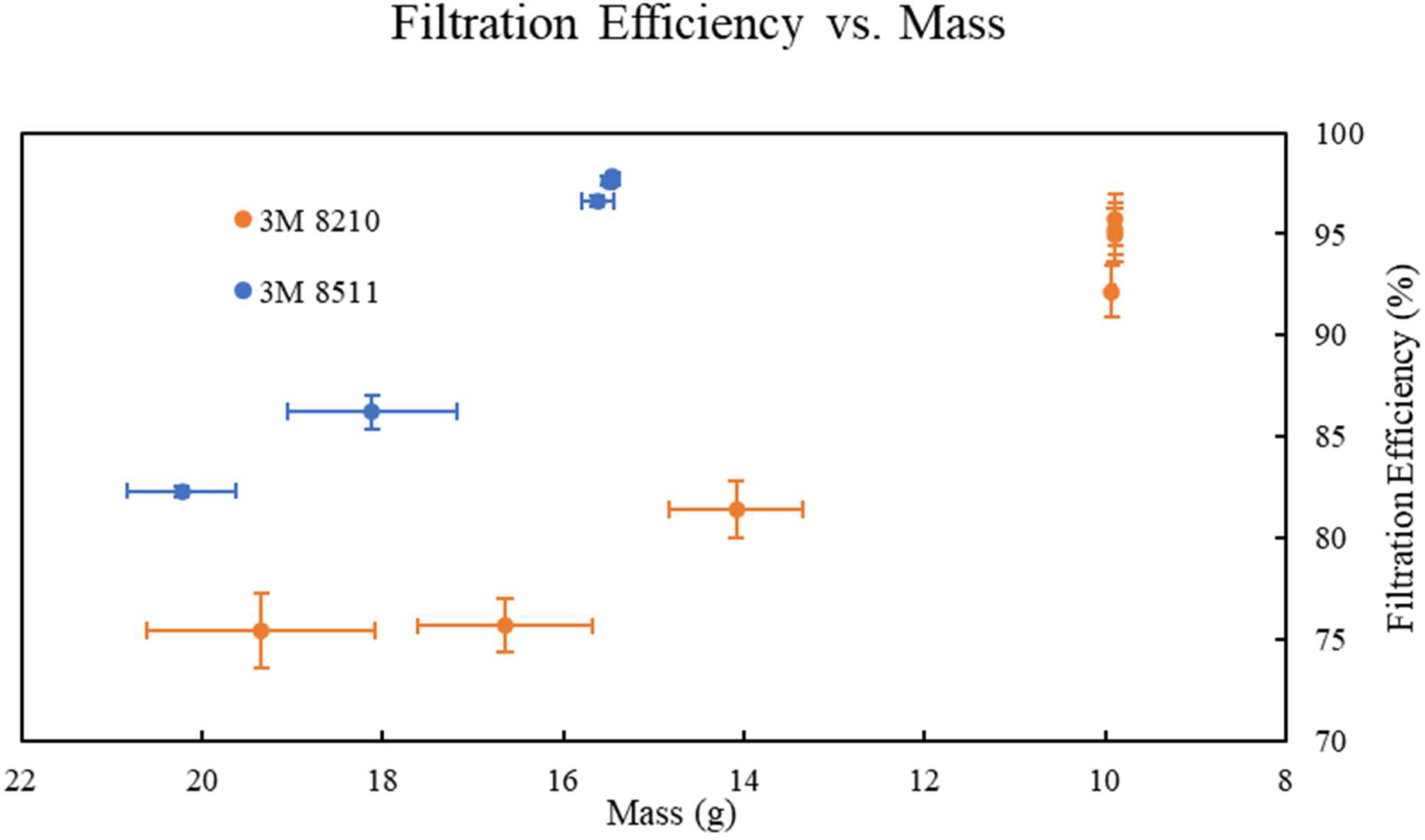
Filtration efficiency vs. mass for two N95 respirators made by the 3M Corporation, models 8210 and 8511. These were soaked in 70% ethanol solution, then allowed to air dry for 2.5 hours until the weight had stabilized. The original dry weights of the masks were 9.88g and 15.46g, respectively. Soaking and then air drying increased the weight of the masks to ∼20.6g and ∼22.8g, respectively. The masks were then pumped on and removed at intervals at specific pressures shown in Figure 3 to record the masks’ mass and efficiency. As the mass of water in the mask decreased, the mask’s efficiency increased along the trend indicated by the colored arrows.

### Optical Microscopy

There are four individual layers of polypropylene microfibers in a 3M 8210 respirator. A 5×5 mm section of one of these layers was isolated and mounted on a glass slide under a thin cover slip. Microscopic images of transmitted light were made with a Leica DM2500 P microscope with a Leica DFC290 HD digital camera. The polypropylene fibers were examined initially dry, then immediately after complete wetting with a 70% ethanol/water solution that was wicked in from the edge. Following this, images were taken periodically during the following 90 minutes when most of the drying was observed to happen. The illumination intensity was not altered during this time so as to preserve relative variations in opacity during the wetting/drying process.

## Results

Table 1 lists some of the medical masks and respirators that we have examined so far. New N95 respirators from 3M typically gave filtration readings above 95% (consistent with their N95 rating from NIOSH), and other mask types with lower ratings gave similarly consistent results. Repeated measurements of the same mask also showed low variance. Although we did not use standard ASHRAE aerosol procedures, data from the experimental setup, shown in Fig. 1, are in agreement with values expected from NIOSH-certified masks. Similarly, the N95 respirators dropped in efficiency by 20-30% following alcohol cleaning; this is consistent with previously reported results (6, 11). Ultimately, we are not relying on the absolute readings, but on the relative changes in filtration efficiency across treatments.

**Table 1:**
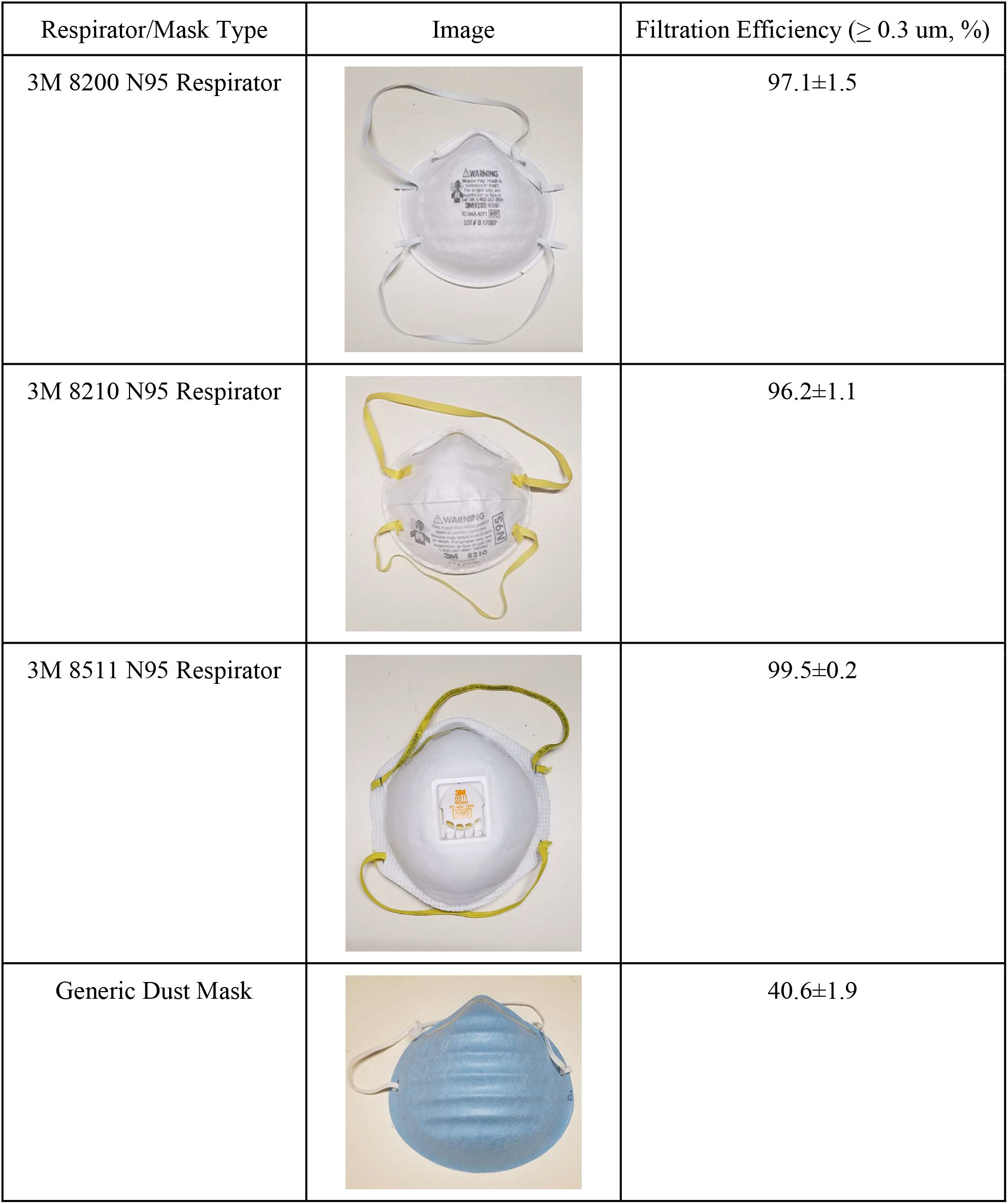
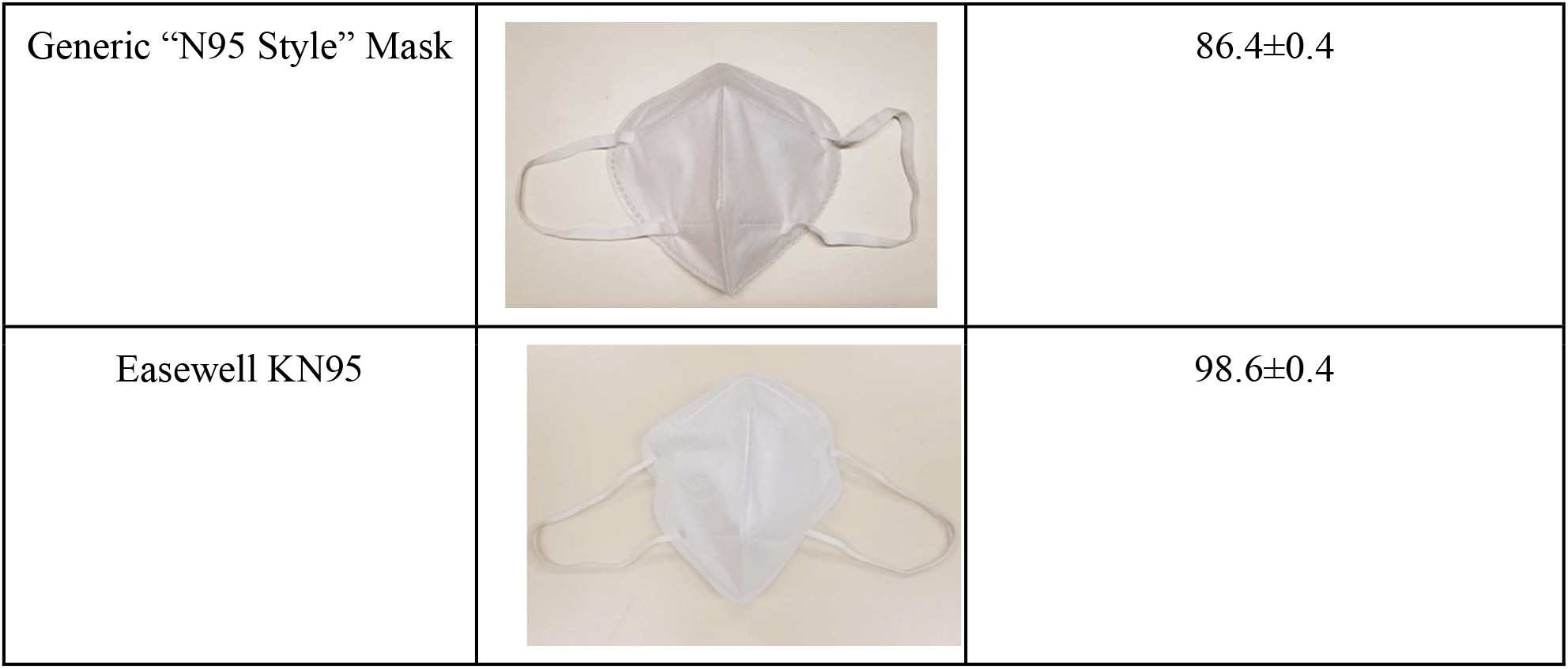
Initial measured filtration efficiency of different respirator / mask types.

We successfully repeated this regeneration procedure on three different types of certified N95 respirators, and one with a KN95 certification, for five cleaning cycles each and saw no long-term degradation (Table 2). After these cycles, all masks remained at ≥95% filtration efficiency. As of this writing, we have conducted twenty-five wet/dry/vacuum cycles on nine different masks, all of which drop in performance after being soaked in 70% ethanol but return to within 99% of their initial filtration efficiency following the vacuum drying step. Data for the effect of mass loss on filtration efficiency for two of the 3M N95 respirators are shown on Fig. 2. The large error in the mass measurements during the main drying interval was due to continued evaporation during the time the masks were on the testing rig with air being forced through them. In both cases, the filtration efficiencies approached their initial values as the mask weights approached their initial values, indicating that the filtration efficiency was being reduced by liquid adsorbed in the washing process.

**Table 2:** Filtration efficiency before and after 5 cycles of washing and vacuum-drying. Initial results are listed from disinfection with 70% v/v ethanol followed by vacuum drying to pressures below 6 mbar. None of the masks dropped below 95% efficiency over the five cleaning cycles, suggesting that these mask types might be used at least 6 times.

| Respirator/Mask Type | % Initial Filtration Efficiency | % Efficiency after 5 Wash Cycles |
| --- | --- | --- |
| 3M 8200 Respirator | 97.1±1.5 | 95.2±1.3 |
| 3M 8210 Respirator | 96.2±1.1 | 97.0±0.2 |
| 3M 8511 Respirator | 99.5±0.2 | 97.2±0.2 |
| Easewell KN95 | 98.6±0.4 | 98.9±0.4 |

Direct measurements of filtration efficiency, as a function of minimum exposed vapor pressure in the vacuum drying step, for these same masks are shown in Fig. 3. In both cases, the major change in filtration efficiency occurred as the pressure was reduced between 13 and 6 mbar, after which the efficiency of both masks returned close to the initial measurements. This range is compatible with the removal of adsorbed water molecules, as considered in the Discussion section below. An additional experiment on one 3M 8200 respirator soaked in pure DI water showed a similar drop in efficiency followed by full recovery to 96.5±0.2% after vacuum drying, supporting the bound water hypothesis.

**Figure 3:**
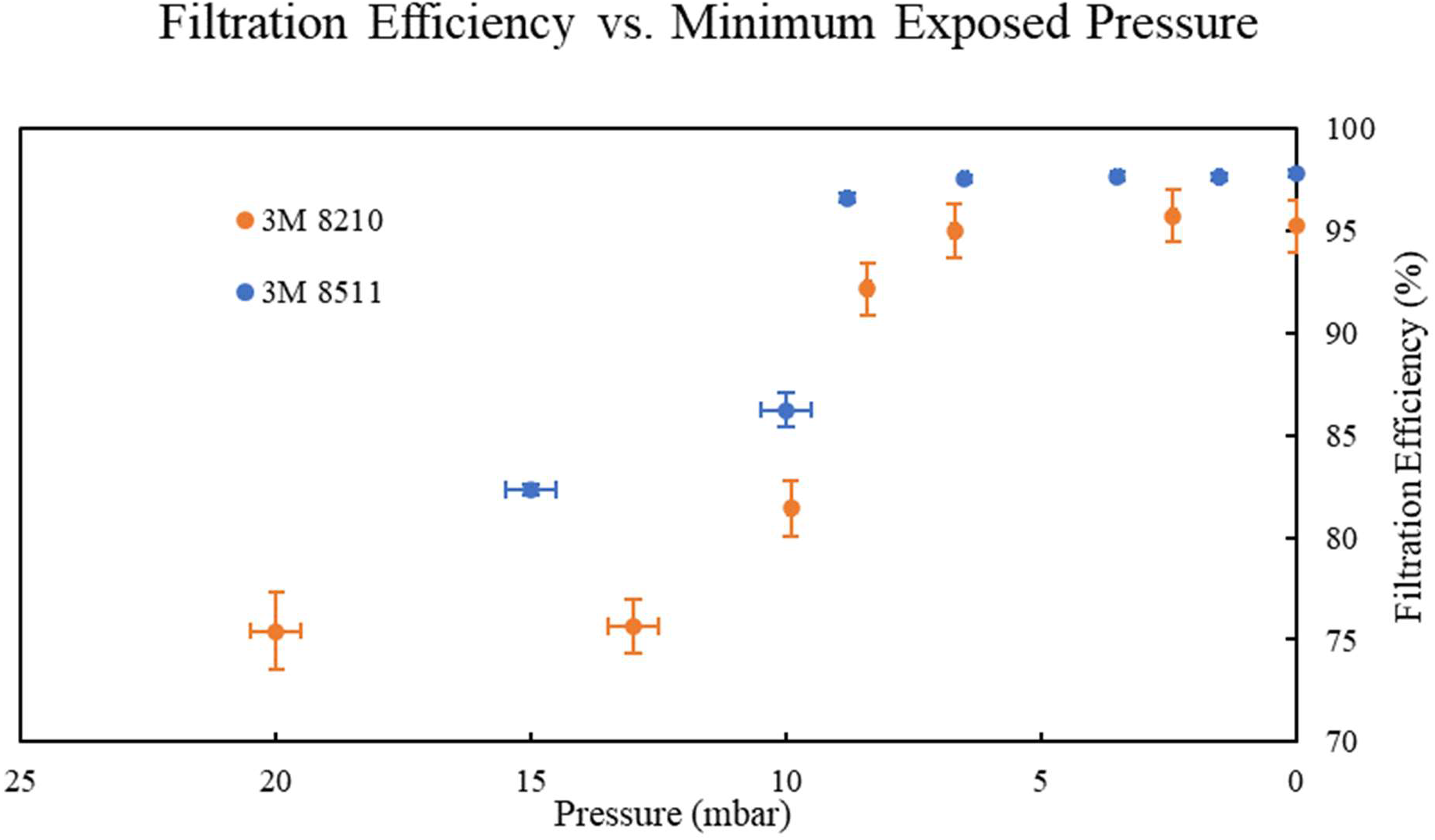
Filtration efficiency vs. minimum exposed pressure for two 3M N95 respirators. Filtration efficiency rapidly increases as pressure drops below ∼13 mbar, corresponding with the removal of water molecules adsorbed to the surface of the microfibers. Vertical error bars are 1 standard deviation, and horizontal error bars represent measurement resolution at the given pressures. Some errors for pressures below 10 mbar are smaller than the symbol size.

Two generic masks of unknown composition and filter efficiency, one of which was labelled as “N95 style” on Amazon, were not measured as having filter efficiency comparable to NIOSH rated N95 masks. This underscores the need for more distributed mask testing capacity as an increasing number of respirators and masks with untested characteristics are brought into medical and professional use.

Optical microscopy of a single microfiber filter layer documented the wetting and drying process (Fig 4). In the dry state (Fig. 4A), polypropylene microfiber bundles are clearly visible as dark, linear features. This is due to the large difference in refractive index between polypropylene and air (1.49 vs. 1.00). Upon addition of the 70% ethanol solution, the intensity of transmitted light increases dramatically through the fibers. This indicates surface wetting of the fibers, which results in index matching at the interface--the smaller difference in refractive indices of polypropylene and 70% ethanol (1.49 vs. 1.37) leads to less scattering of light. During the progressive drying process, the transmitted light intensity decreases as expected, but in an irregular fashion. Bubbles develop and major bundles of fibers remain bright (wet) until they dry at the very end. We observe no significant motion or displacement of the majority of the fibers during this wet/dry cycling process.

**Figure 4:**
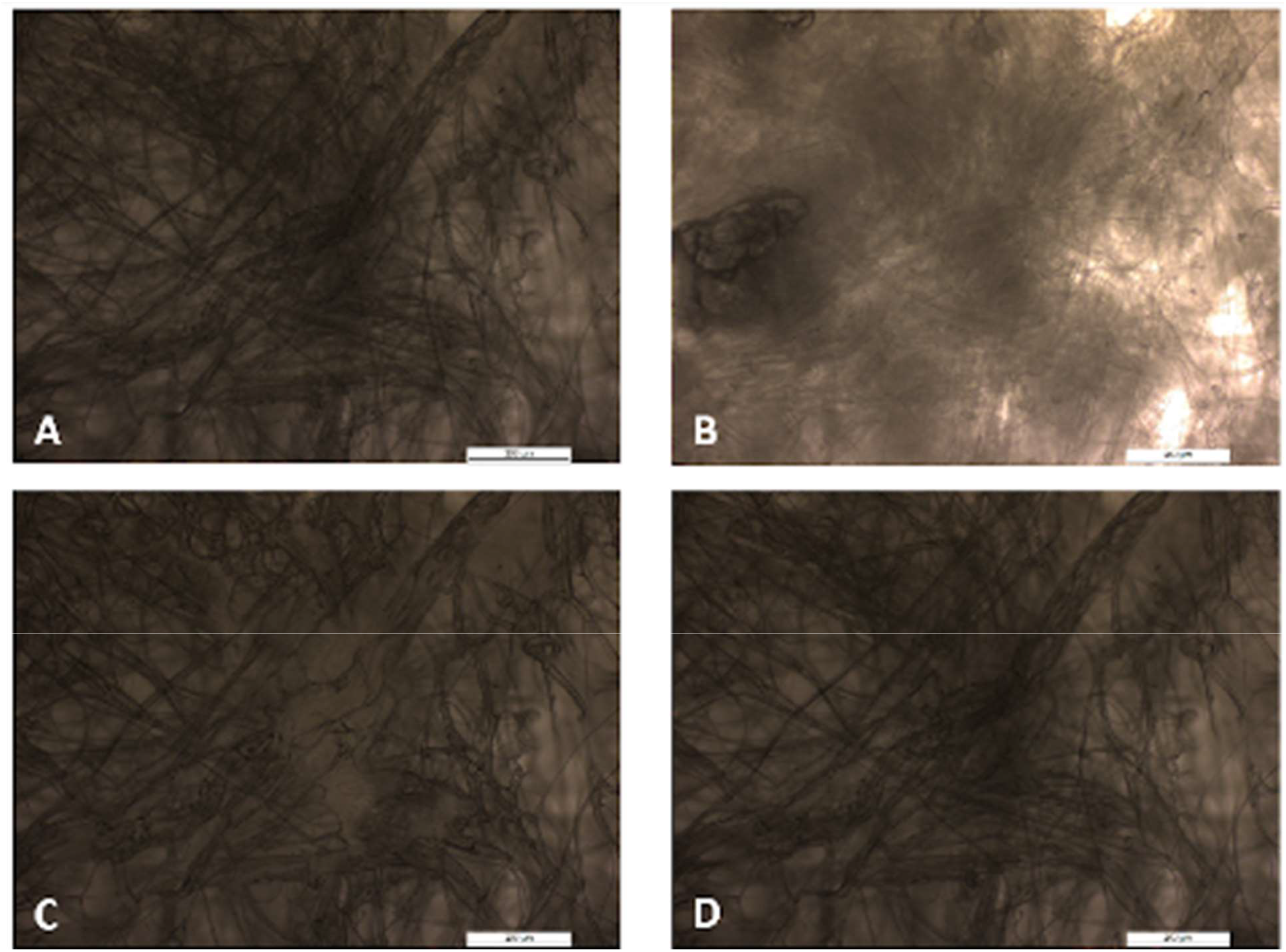
Optical Images of a polypropylene microfiber filter from a 3M 8210 mask, mounted on an aluminum plate for imaging and filtration tests. A) Initial filter prior to any treatment. B) Filter after saturation in 70% ethanol. Transmitted light increases, due to surface wetting and closer matching of the refractive indices between the 70% ethanol and the polypropylene fibers. C) 45 minutes after soaking. The solution is now only present in the areas of high fiber density. D) 84 minutes after soaking. This image is nearly identical to that on A), indicating that the filter has dried. Scale bars in all images are 200 µm.

## Discussion

### Proposed Mechanism of Action

Previous studies have proposed that the observed drop in filtration efficiency is due to ethanol permanently neutralizing surface charges on the polypropylene microfibers (5, 9, 11). Our data point to a different mechanism: water remains adsorbed onto the fiber surfaces, and vacuum drying then removes those layers. The pressure at which we observed filtration performance recover is well below the vapor pressures where ethanol and bulk water would normally be removed (58 & 23 mbar at 20°C, respectively). Instead, the performance recovers sharply in a pressure range corresponding to the removal of water molecules that are adsorbed to surfaces (1-20 mbar) (13).

This surface wetting mechanism for performance degradation is consistent with the nature of the mask materials. The active material of the 3M N95 masks is melt-blown, polypropylene microfibers containing embedded, static charges (9, 11). SEM images indicate that the diameters of the fibrous strands are on the order of ∼1 μm (14), giving the filter a huge specific surface area. When the fibers are soaked with the ethanol solution and air dried, thin liquid films could be left on the material. Assuming the diameter of the polypropylene fibers to be ∼1 μm, a monolayer of water (∼0.1 nm thick) would increase the mass of a mask by around 25% (7). This is consistent with our observations: when a mask is treated and left to air dry overnight, a residual of several grams is left.

As the solution dries, the mixture evaporates with ethanol preferentially being in the vapor. The result of several hours of air drying is a film of primarily water deposited on the surface of the fibers. Our microscopy images indicate that the final fraction of water, while not contributing a large amount of mass, is wicked into the dense networks of microfibers. These networks would normally have a large internal surface area available for particle filtration interactions. But having water wicked into the network decreases the surface area available and reduces filtration efficiency. Furthermore, the low surface area available also inhibits evaporation of the water stuck between fibers. We removed this water by drying the mask under vacuum, to decrease the partial pressure of water and increase the mean free path of water molecules.

Our results indicate that mask performance scales with how well the masks are dried, as measured both by the mass of the masks and the vacuum level used to dry them. A mask soaked in 70% v/v ethanol and dried in air overnight weighs 2-4 g more than it did originally and has substantially reduced filtration efficiency (Fig. 2). Furthermore, our data show that the filtration efficiency scales inversely with the amount of water adsorbed. Vacuum treatment of masks at pressures between 10 and 6 mbar restored the filtering efficiency (Fig. 3).

Our results do not rule out the operation of the previously proposed ethanol-based electret discharge mechanism, it may coexist with the water-based adsorption mechanism proposed here. Our measurements suggest that the water-based mechanism accounts for the majority of the observed loss of filtration efficiency, at least within our testing regime.

### Relationship to other tests and studies

Previous studies of ethanol-based disinfection used a variety of methods for disinfection and filtration measurement. Ethanol concentrations ranged from 70-75%, and drying protocols were not described (6, 11). Filtrations measurements were done either on filter material sealed to a NIOSH-standard testing rig (11), or on a mask undergoing quantitative fit testing (6). Our experiments indicate that drying protocols may have a major effect on results, but other differences between our experiments may also have incremental effects.

Further testing will be necessary to determine how treated respirators perform under the more challenging conditions contained in the NIOSH standards. Such tests will be required to determine whether the respirators continue to meet the N95 standard after treatment. Under higher airflow rates, and high loads of neutral aerosol standards, overall filtration efficiencies may drop, and the relative contributions of discharge and adsorption mechanisms may shift. That said, the mild filtration challenges in our tests produced clearly measurable efficiency losses, suggesting that these water adsorption effects would be substantial under the stronger NIOSH challenges. In addition, it is obvious that masks that do poorly on our tests (e.g., the ‘generic’ masks shown on Table 1) are allowing a high enough fraction of particles to penetrate them such that they ought to fail the more rigorous NIOSH standards; for this reason simple testing rigs can be useful to weed out the bad masks before bulk purchases are made.

The test setup we use is analogous to the quantitative fit tests used to assess how well a given mask conforms to a given individuals’ face. This is in contrast to the testing rigs used to certify masks, which test the performance of the masks’ filtration material by sealing the mask onto the flat surface of the device. Running such filtration medium tests on our setup involves replacing the head model with a flat plate attachment. As our tests used a head model, it is possible that fit factor may have contributed to our results. Our error analysis suggests that the mask fit was at least consistent across the repeated measurements contributing to a single data point. But this does not rule out systematic changes in fit across different stages of drying.

### Limitations and Caveats

We avoided the use of tap water in our alcohol mixture because of the previous suggestions of interactions with surface charges on the microfibers. Although our findings support surface tension mechanisms instead, we do not know if ions in tap water have an effect. This remains to be tested.

It is not known how generalizable these results are across other types of masks or similar decontamination procedures. For example, both ethanol and isopropanol are known to be effective decontaminants, but whether our method extends to isopropanol is unknown. Unlike ethanol, isopropyl alcohol damages polyester (8) and may degrade other components of the masks and respirators.

The procedure outlined here for regenerating mask filtering efficiency has not been approved by the FDA, NIOSH, or any other relevant regulatory agencies, although the use of the alcohol solution is a well-vetted technique for disinfection (3). Our results indicate that ethanol-based disinfection should not be ruled out in protocol development.

Our testing station tests filtering performance on particles in ambient air, rather than ASHRAE aerosol standards. However, our particle counter directly measures particle sizes, based on calibrations that are NIST-referenced and meet or exceed CE and ISO 21501 certification. The distribution of particle sizes was appropriate, with ∼90% of particles falling in the 0.3-0.5 µm bin, and our analyses were based on data from that bin. Further, our tests produced the expected results on certified masks, and replicated the results from other experiments using NIOSH-defined, standard methods. Ultimately, we would like to determine the relationship between measurements from inexpensive, accessible designs and measurements that meet the N95 respirator NIOSH testing standards. The NIOSH test guidelines (15) mandate specific particle size distributions and air flow rates for salt or DOP aerosols (e.g. for salt aerosols, a median particle diameter of 0.075 ± 0.020 µm and a standard geometric deviation not exceeding 1.86 at the specified test conditions as determined with a scanning mobility particle sizer or equivalent, and an air flow rate across the respirators of 85 ± 4 L/min). Our current mask test system does not meet these more rigorous specifications.

### Implications

Our results indicate that water adsorption can cause loss of filtration efficiency, even in respirators that seem dry to the touch. This may account for some of the losses in efficiency seen in previous tests of ethanol disinfection. Further testing under NIOSH standards is needed to determine filtration levels after respirators are completely dry, and to see if safe ethanol-based protocols can be designed.

Although adsorbed water causes a loss of filtration efficiency, it is not the case that the introduction of water into a mask automatically leads to filtration loss. Liao et al. (11) found that filtration was maintained across several cycles of steam treatment, until a sudden drop after the fourth cycle. We have made similar informal observations, finding that filtration efficiency remains relatively constant even after saturated water vapor exposure has increased the mass of the mask by several grams. This indicates that mere *absorption* of moisture by the mask is not sufficient to decrease performance. Some critical conditions are required for surface *adsorption* to occur, and for filtration to decline.

Determination of those conditions will be important for recognizing the contexts in which one should guard against risk. Incidental sources of moisture such as rain, spills, or perspiration might also affect filtration if accumulated in sufficient amounts. While current clinical and materials safety guidelines already advise against use of damp respirators (1), it is not easy to detect the amounts of residual adsorbed water that will degrade filtration efficiency. Thus, periodic drying practices may be advised as part of reconditioning protocols, regardless of the chosen disinfection technique.

Our ethanol and vacuum-drying experiment results suggest that the surface wetting mechanism may be generalizable in two directions. First, some of the other aqueous disinfectants previously suspected of chemically damaging microfiber filters may in fact be viable with proper drying. Second, alternative drying methods known to remove surface-adsorbed water (e.g., dry gases) are also likely to restore filtration. Given enough time, conventional drying methods may also work. Earlier literature includes several tests of other aqueous disinfectants, in which filtration was found to be intact after either 16+ hours of forced-air drying (2) or 72 hours of hang drying (16). The more recent manuscripts that reported significant deterioration in the efficacy of the respirators did not specify the air-drying times used in their tests (6, 11). Future research and regulatory guidance on reconditioning protocols should include specific drying methods in their considerations.

## Data Availability

All data are available upon request.

## Acknowledgments

We thank Mr. Masamoto Horikawa, Mr. Hironori Hidaka and Dr. Atsuko Kobayashi of the Tokyo Institute of Technology for the early version of the control software for the laser particle counter.

